# SARS-CoV-2 infects blood monocytes to activate NLRP3 and AIM2 inflammasomes, pyroptosis and cytokine release

**DOI:** 10.1101/2021.03.06.21252796

**Authors:** Caroline Junqueira, Ângela Crespo, Shahin Ranjbar, Jacob Ingber, Blair Parry, Sagi Ravid, Luna B. de Lacerda, Mercedes Lewandrowski, Sarah Clark, Felicia Ho, Setu M. Vora, Valerie Leger, Caroline Beakes, Justin Margolin, Nicole Russell, Lee Gehrke, Upasana Das Adhikari, Lauren Henderson, Erin Janssen, Douglas Kwon, Chris Sander, Jonathan Abraham, Michael Filbin, Marcia B. Goldberg, Hao Wu, Gautam Mehta, Steven Bell, Anne E. Goldfeld, Judy Lieberman

## Abstract

SARS-CoV-2 causes acute respiratory distress that can progress to multiorgan failure and death in some patients. Although severe COVID-19 disease is linked to exuberant inflammation, how SARS-CoV-2 triggers inflammation is not understood. Monocytes are sentinel blood cells that sense invasive infection to form inflammasomes that activate caspase-1 and gasdermin D (GSDMD) pores, leading to inflammatory death (pyroptosis) and processing and release of IL-1 family cytokines, potent inflammatory mediators. Here we show that ~10% of blood monocytes in COVID-19 patients are dying and infected with SARS-CoV-2. Monocyte infection, which depends on antiviral antibodies, activates NLRP3 and AIM2 inflammasomes, caspase-1 and GSDMD cleavage and relocalization. Signs of pyroptosis (IL-1 family cytokines, LDH) in the plasma correlate with development of severe disease. Moreover, expression quantitative trait loci (eQTLs) linked to higher *GSDMD* expression increase the risk of severe COVID-19 disease (odds ratio, 1.3, p<0.005). These findings taken together suggest that antibody-mediated SARS-CoV-2 infection of monocytes triggers inflammation that contributes to severe COVID-19 disease pathogenesis.

**One sentence summary:** Antibody-mediated SARS-CoV-2 infection of monocytes activates inflammation and cytokine release.

---

In a small subset of mostly elderly patients or patients with comorbidities, SARS-CoV-2 causes severe COVID-19 disease marked by acute respiratory distress that can progress to multiorgan failure and death (*1*). Severe disease is linked to an overly exuberant inflammatory response, including elevated serum pro-inflammatory cytokines, C-reactive protein, and lactate dehydrogenase (LDH) (*2–6*). Increased chronic inflammation is associated with aging (“inflammaging”) and the comorbidities linked to severe COVID-19 disease (*7*). Myeloid cells (monocytes, macrophages, dendritic cells) are sentinels that sound the innate immune alarm by sensing invasive infection and danger to activate inflammasomes (*8*). They are usually the most important source of inflammatory cytokines during inflammation, and their activation is required to process and release IL-1 family cytokines, arguably the most potent inflammatory mediators in the body (*9*). However other pathways, including NF-κB activation by Toll-like receptors or the TNF receptor superfamily and TH17 lymphocyte cytokines, can also cause severe inflammation.

Because inflammasome activation in myeloid cells is a major mediator of inflammation (*2, 10*), we examined blood of SARS-CoV-2-infected donors for signs of inflammasome activation. When inflammasomes sense danger or infection, they recruit the ASC adaptor and assemble into large supramolecular complexes that recruit and activate the inflammatory caspase-1, which in turn processes interleukin (IL)-1 family pro-cytokines and the pore-forming protein GSDMD that damages the cell membrane, leading to cell death and inflammatory cytokine release (*8*). Cell membrane rupture during pyroptosis releases large proteins such as the tetramer LDH (144 kDa), a pathognomonic feature of pyroptosis (*8*).

But what activates inflammasomes in COVID-19 patient monocytes? Since inflammasomes sense invasive infection, one possibility is that they might be infected. However, monocytes are generally thought not to express ACE-2, the viral receptor for entry. Indeed, we found that healthy donor (HD) monocytes do not express ACE-2 even when they are activated. However, ACE-2 may be dimly expressed on COVID-19 patient monocytes and a recent paper suggests that tissue macrophages might be infectable (*11*). Indeed, we found that about 10% of freshly isolated monocytes from COVID-19 patients were infected and replicating the virus and the infected cells had activated inflammasomes. Thus, monocyte infection activates inflammasomes. However, HD monocytes were not susceptible to SARS-CoV-2, but infection was significantly increased after monocytes were activated and the virus was preincubated with SARS-CoV-2 monoclonal antibodies or patient plasma, but not with antibody-depleted plasma. suggesting that inflammation and antibodies present in infected patients increase monocyte susceptibility.

## Circulating monocytes and plasma of COVID-19 patients show signs of pyroptosis

To analyze cell death, freshly isolated mononuclear cells from 19 healthy donors (HD) and 22 COVID-19 patients seen in the emergency department (ED) of Massachusetts General Hospital were stained for hematopoietic cell markers, a small fixable dye (Zombie Yellow) that enters dying cells whose plasma membrane is damaged and annexin V, which identifies cells undergoing programmed cell death (Fig. 1a-c, Fig. S1a, Table S1). Although annexin V+Zombie-apoptotic cells did not increase in any subpopulation in COVID-19 samples, on average ~11% of monocytes of COVID-19 patients took up Zombie dye, a sign of ongoing membrane damage consistent with pyroptosis. Consistent with previous reports (*12*), a reduced proportion of CD14+CD16-classical monocytes and more CD14+CD16+ intermediate monocytes were observed in COVID-19 patients compared to HD (Fig S1b, c). The intermediate state is a sign of monocyte activation (*12*).

**Figure 1.**
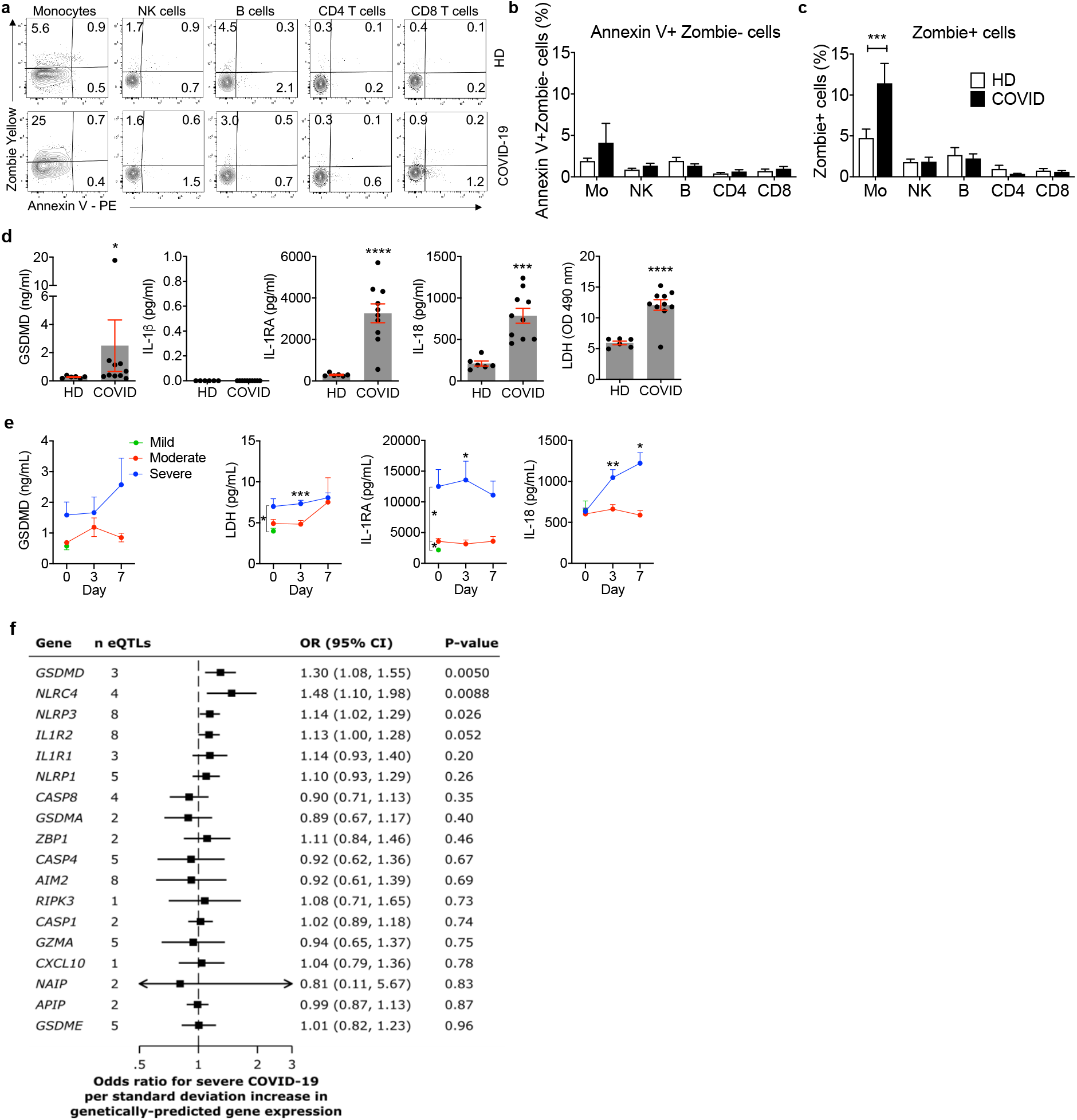
Circulating monocytes from COVID-19 patients are undergoing pyroptosis a-c, Representative flow cytometry contour plots (**a**) and percentage of lymphocyte and monocyte subsets staining for Annexin V only (**b**) or Zombie (**c**) in 19 healthy donors (HD) and 22 SARS-CoV-2 infected patients. **d**, Concentration of gasdermin D (GSDMD), pyroptosis-related cytokines (IL-1β, IL-1RA, IL-18) and lactate dehydrogenase (LDH) activity in the plasma of HD (n=6) or SARS-CoV-2 positive patients (COVID, n=10). **e**, Plasma pyroptosis biomarkers GSDMD, LDH activity, IL-1RA and IL-18 at presentation and during hospitalization in COVID-19 patients with mild, moderate and severe COVID-19 acuity scores (n=60). **f**, Odds ratio for severe COVID-19 disease relative to eQTLs that are associated with increased gene expression of pyroptosis, necroptosis and death receptor-related genes. Data show mean ± S.E.M or odds ratio (95% confidence interval). *p<0.05, **p<0.01, *** p<0.001, ****p<0.0001 by two-way ANOVA (b,c) Mann-Whitney or Kolmogorov-Smirnov test (d) and multiple t-tests (e).

Many studies have reported signs of inflammation in the plasma of COVID-19 patients and some have reported increased plasma biomarkers specifically linked to canonical inflammasome and gasdermin activation, such as IL-1 family cytokines and LDH, which is only released in necrotic forms of cell death, including pyroptosis (*13–15*). Elevated LDH is one of the best correlates of severe COVID-19 disease (*6*). However, none have looked at plasma GSDMD, which is selectively released during pyroptosis (*16*). Plasma from the HD and COVID-19 patients evaluated above was assessed by multiplex ELISA and LDH activity assay for specific inflammatory markers of pyroptosis (GSDMD, IL-1β, IL-1RA, IL-18, LDH) (Fig. 1d) and for other markers of inflammation that are not pyroptosis-specific (inflammatory cytokines IL-6, TNFα, IL-17/17A; growth factors IL-7, G-CSF; chemokines CCL7, CXCL9, CXCL10) and for interferons (IFNβ, IFNγ) (Fig. S 2a). All of the specific pyroptosis markers, except IL-1β, were significantly elevated in COVID-19 patient plasma compared to HD. Plasma IL-1β was below the level of detection in all samples, which was not surprising since it is rapidly cleared from the blood and usually not detected even in patients with severe disease caused by ongoing pyroptosis, such as those bearing constitutively active *NLRP3* mutations (*9*) or with IL-1-mediated systemic juvenile idiopathic arthritis (*17*). However, its antagonist IL-1RA, which can be used as a surrogate (*9*), was greatly increased in COVID-19 samples. IL-6 and TNFα, which can be secondarily increased following IL-1 family cytokine binding to IL-1 receptors, and many of the growth factors and chemokines measured were also highly elevated in COVID-19 plasma compared to HD, as has been reported (*13*). In contrast, the interferons were not detected in the plasma, consistent with previous observations that SARS-CoV-2 infection inhibits the interferon response (*18, 19*). We also did not observe increased IL-17 levels. Taken together, these results provide evidence of ongoing pyroptosis in blood monocytes and plasma of COVID-19 patients.

### Plasma markers of pyroptosis correlate with severe COVID-19 disease

To determine if pyroptosis-related biomarkers correlate with COVID-19 disease severity, plasma from 60 COVID-19 patients who presented to the ED were analyzed for GSDMD, LDH and plasma cytokines at presentation and on days 3 and 7 for those patients who were hospitalized (Fig. 1e, Fig. S2b, Table S2). Patients were grouped into mild, moderate or severe disease using the MGH COVID Acuity scale (*20, 21*). Moderate disease was defined as requiring supplemental O2 during hospitalization and severe disease was defined as requiring mechanical ventilation and/or causing death. Plasma levels of the pyroptosis-specific markers, GSDMD, LDH, IL-1RA and IL-18, were all elevated in severe patient samples compared to those with mild or moderate disease, but the increase in GSDMD did not reach significance (Fig. 1e). Other markers of inflammation that are not specific for pyroptosis (C-reactive protein, D-dimer, IL-17, CCL7, CXCL10) were significantly increased early during hospitalization in patients who developed severe COVID-19, but others (IL-6, TNFα, CXCL9) were not in our samples (Fig. S2b). Although GSDMD levels did not significantly correlate with disease severity, GSDMD plasma levels measured in the ED correlated significantly with a marker of pyroptosis (IL-1RA) and with some other markers of inflammation (IL-6, G-CSF, CXCL10) (Fig. S2c). Furthermore, plasma IL-1RA also significantly correlated with plasma IL-6, TNFα, G-CSF, and CXCL10 on day 0 (Fig. S2d). Thus, pyroptosis biomarkers at presentation correlate with COVID-19 disease severity.

### *GSDMD* eQTLs increase the risk of severe COVID-19 infection

To probe further whether pyroptosis might be associated with severe COVID-19 infection, we used Mendelian randomization analyses to examine whether expression quantitative trait loci (eQTLs) linked in eQTLGen (*22*) to increased blood expression of 18 immune genes are associated with severe COVID-19 disease. The gene products of the genes analyzed mostly play roles in inflammation and/or programmed necrosis - 11 inflammasome and pyroptosis-related genes: *AIM2, NLRC4, NLRP1, NLRP3, NAIP, CASP1, CASP4, GSDMA, GSDMD, GSDME, GZMA;* 3 genes related to inflammatory cytokine/chemokine signaling: *IL1R1, IL1R2, CXCL10;* 2 necroptosis (another inflammatory programmed necrosis pathway) genes: *RIPK3, ZBP1;* and 2 death receptor or apoptosis signaling genes: *CASP8, APIP*. Data were from case-control study cohorts of the COVID-19 Host Genetics Initiative release #4 (October, 2020) (*23*) (Fig. 1f, Table S3). In the analysis, the contribution of each eQTL for a given gene was weighted according to its prevalence in the general population. Comparing 4336 severe COVID-19 cases with 623,902 population controls, *GSDMD* eQTLs (of which there are 3 (Fig. S3a)) were most significantly associated with increased COVID-19 respiratory failure (odds ratio 1.30, 95% confidence interval (1.08, 1.55), p<0.005). eQTLs associated with higher expression of two inflammasome genes (*NLRC4* and *NLRP3*) were also significantly linked to severe COVID-19 disease. The odds ratio for the 4 *NLRC4* eQTLs was 1.48 (95% confidence interval (1.10, 1.98), p<0.008) and for the 8 *NLRP3* eQTLs was 1.14 (95% confidence interval (1.02, 1.29), p<0.03). A parallel analysis of eQTL links to COVID-19 hospitalization (6406 hospitalized cases versus 902,088 population controls, Fig. S3b and Table S3) identified only 1 significant association with *AIM2*, another inflammasome gene, which has 8 eQTLs. However, in this case *AIM2* eQTLs associated with increased COVID-19 hospitalization led to a lower risk of COVID-19 hospitalization (odds ratio 0.77, 95% confidence interval (0.61,0.97), p<0.03). When a smaller dataset compared COVID-19 patients with severe vs mild disease (269 cases of severe COVID-19 with 688 controls of non-hospitalized COVID-19 patients) (Fig. S3c and Table S3), patients with severe COVID-19 disease had significantly more eQTLs linked to higher expression of only one gene, the gasdermin *GSDME* with 5 eQTLs (odds ratio 1.93, 95% confidence interval (1.15, 1.3.24), p<0.013). In all of these analyses, eQTLs for none of the 7 analyzed genes that were not directly associated with inflammasome activation and pyroptosis were significantly enriched in patients with more severe COVID disease. Thus, the genetic link between increased gasdermin and inflammasome eQTLs and severe COVID-19 infection further supports a role for pyroptosis in clinical deterioration. The stronger genetic link between gasdermins and inflammasomes in severe COVID disease than in COVID-19 hospitalized patients suggested by our analysis (despite the larger sample size for the hospitalized patient analysis), hints that pyroptosis may be especially important in the immunopathogenesis that accompanies the transition from initial pneumonitis to respiratory failure and systemic disease.

### Circulating monocytes have activated NLRP3 and AIM2 inflammasomes

The above data suggested that circulating monocytes in COVID-19 patients might die of pyroptosis and release inflammatory cytokines and cause cytokine storm and contribute to poor outcome. There are 27 potential human canonical inflammasome sensors (22 NOD-like receptors (NLR), 4 AIM2-like receptors (ALR) and pyrin), but not much is known about how viruses interact with them (*8*). The NLRP3 inflammasome, which detects K^+^ efflux generated by a variety of stimuli including extracellular ATP, bacterial toxins or disruption of the cell membrane, could be activated by lytic SARS-CoV-2 infection itself or by specific viral proteins (*24, 25*). Three SARS-CoV-2 proteins, Orf3a, Orf8b and the E envelope, are “viroporins” (ion channels) that activate NLRP3 by K^+^ efflux when ectopically expressed (*26–29*). Orf3 and Orf8 are encoded by the pathogenic, but not the avirulent, human CoVs, suggesting that NLRP3 activation might differentiate virulent from avirulent coronavirus infection. Interestingly, bats, the natural zoonotic hosts of SARS-CoV and SARS-CoV-2, have a dampened NLRP3 response to infection with multiple viruses, including MERS-CoV, which might explain their ability to tolerate these infections despite high viral loads (*30*). To probe whether circulating monocytes from COVID-19 patients are undergoing pyroptosis, freshly isolated, purified monocytes from 3-8 HD or COVID-19 patients were stained for the common inflammasome adapter ASC, activated caspase-1 (visualized by fluorochrome-labeled inhibitor of caspases assay (FLICA)) and GSDMD and analyzed by imaging flow cytometry for their expression and intracellular distribution (Fig. 2 and Fig. S4). When a canonical inflammasome is activated, a large micron-sized inflammasome-ASC-caspase-1 speck is formed (*8*). A significant proportion of freshly isolated monocytes from COVID-19 patients, compared to HD, showed activated caspase-1 and ASC specks (Fig. 2a-c), confirming that circulating monocytes in COVID-19 patients are spontaneously undergoing pyroptosis. Most of the cells with ASC specks (>80%) also had co-localized activated caspase-1 specks, visualized by fluorochrome-labeled inhibitor of caspases (FLICA) assay (Fig. 2d). When these monocytes were treated with a low concentration of nigericin (20 µM) for 30 minutes, which triggers the NLRP3 inflammasome, all the samples had some cells with ASC-caspase-1 specks, but in the COVID-19 samples these were increased approximately three-fold, indicating that they were more prone to stimulated pyroptosis. As a control, a monocytic cancer cell line (THP-1) was treated with nigericin.

**Figure 2.**
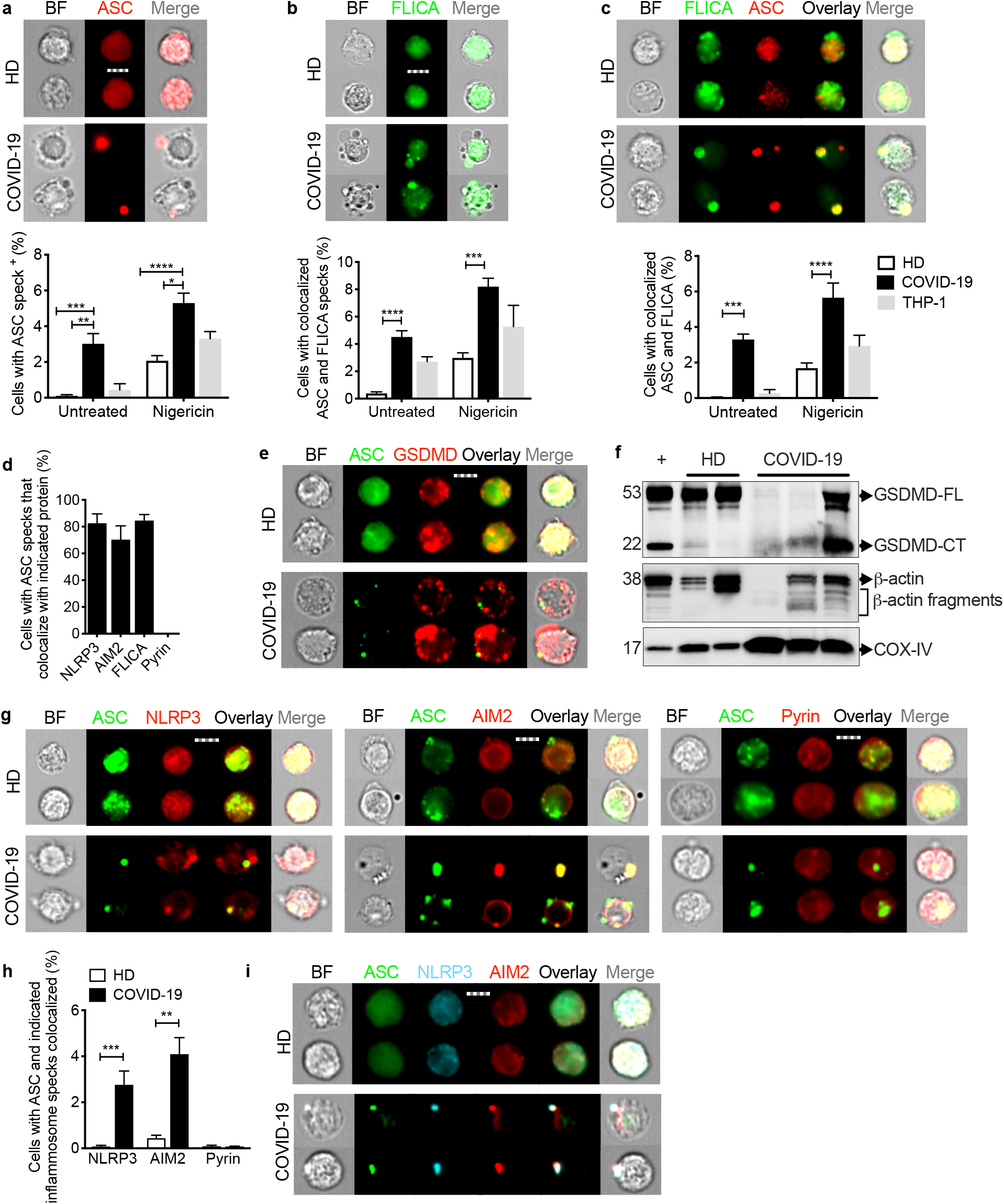
Monocytes from COVID-19 patients have activated inflammasomes, caspase-1 and gasdermin D. Circulating monocytes from healthy donors (HD) or COVID-19 patients were treated or not with nigericin, and then analyzed by imaging flow cytometry for caspase-1 activation (by FLICA assay before fixation) and fixed and stained for the indicated markers. **a-c**, Percentage of monocytes with activated ASC, n=5 (**a**) or caspase-1, n=5 (**b**) or colocalized ASC/caspase-1 specks, n=5 (**c**). Representative images are shown at left and quantification is graphed at right. **d**, Percentage of ASC-speck-containing monocytes with colocalized NLRP3 (n=6), AIM2 (n=4), caspase-1 (n=6) or pyrin (n=4) specks. **e**, Representative images of ASC and GSDMD co-stained monocytes (from 4 independent experiments). f, Immunoblot of lysates of freshly isolated purified HD and COVID-19 monocytes and of HD monocytes treated with LPS and nigericin (+) probed with GSDMD mAb that recognizes full length (GSDMD-FL) and the cleaved C-terminal fragment (GSDMD-CT) (top). Anti-β-actin (middle) and COX-IV (bottom) are loading controls. **g**,**h**, Representative images of ASC co-staining with NLRP3 (left), AIM2 (middle) and pyrin (right) (**g**) and quantification of monocytes showing colocalization of ASC specks with each inflammasome (**h**). **i**, Representative images of costaining of ASC, NLRP3, and AIM2 (from 3 independent experiments). Scale bar, 7 μm. BF, brightfield. Mean ± S.E.M. is shown. *p<0.05, **p<0.01, ***p<0.001, ****p<0.0001 by two-way ANOVA with Tukey’s multiple comparisons test.

As expected GSDMD did not co-localize with ASC-caspase-1 specks in any of the samples (Fig. 2e). GSDMD staining was different in ASC speck+ COVID-19 patient freshly isolated monocytes, compared to HD and to ASC speck-negative COVID-19 patient monocytes – in cells with ASC specks, GSDMD redistributed from mostly cytoplasmic staining to form prominent cell membrane puncta, consistent with GSDMD cleavage and pore formation (Fig. 2e and Figure S4b). Immunoblots of lysates of freshly isolated HD and COVID-19 patient monocytes and of HD monocytes treated with LPS and nigericin to activate the NLRP3 inflammasome were probed for full-length GSDMD (GSDMD-FL) and its cleaved CT fragment (GSDMD-CT) and the housekeeping proteins, β-actin and COX-IV (Fig. 2f). Although GSDMD-FL was observed in all the HD samples, it was only detected in 1 of 3 COVID-19 samples. A prominent GSDMD-CT fragment was detected in LPS + nigericin-treated HD monocytes and in the COVID-19 monocyte sample in which GSDMD-FL was detected and it was also clearly detected in one of the other COVID-19 samples. Although mitochondrial inner membrane-anchored COX-IV was detected in all the samples, FL β-actin was not detected in one of the COVID-19 samples, but immunoreactive β-actin fragments were detected in all the COVID-19 samples and in the NLRP3-activated HD monocytes. During pyroptosis, cleaved GSDMD and actin are released, the actin cytoskeleton disintegrates and pyroptotic cells no longer staining for actin, while membrane-bound proteins, like COX-IV, are mostly retained (*16, 31*). Thus, the imaging flow cytometer and immunoblot data indicate ongoing GSDMD cleavage and pyroptosis in COVID-19 monocytes.

To identify the inflammasome sensor activated in COVID-19 patient monocytes, freshly isolated HD and COVID-19 patient monocytes were costained for ASC and 3 canonical inflammasomes (NLRP3, AIM2 (activated by cytoplasmic DNA) or pyrin (activated by bacterial toxins, as a negative control)) (Fig. 2d, g-i) (*32, 33*). COVID-19 patient monocytes with ASC specks co-localized NLRP3 and AIM2 with ASC specks, but there were no pyrin specks, as expected. Thus, both NLRP3 and AIM2 were activated to form inflammasomes in COVID-19 patient monocytes. The activation of the DNA sensor AIM2 was unexpected, although AIM2 has been shown, in rare cases, to be activated by RNA viruses by an unclear mechanism (*34–36*). AIM2 might also sense host genomic or mitochondrial DNA; notably mitochondrial membranes are reported to be damaged during pyroptosis (*37*). Because assessment of cellular co-localization is more convincing using confocal microscopy than by imaging flow cytometry, Z stacks and 2D projection of confocal microscopy images of HD and COVID-19 patient monocytes co-stained for ASC and activated caspase-1, NLRP3, AIM2 or pyrin inflammasomes were also analyzed. Confocal images confirmed the imaging flow cytometry co-localizations (Fig. S4c). Almost all of the cells with ASC specks had co-localized NLRP3 and AIM2 specks (Fig. 2d) and ASC, NLRP3 and AIM2 co-localized together (Fig. 2i). These data demonstrate that NLRP3- and AIM2-ASC-caspase-1 inflammasomes are activated and GSDMD relocalizes to the cell membrane in a significant fraction of circulating monocytes in COVID-19 patients. Taken together with the large proportion of dying Zombie+annexin V-circulating monocytes we observed (Fig. 1a–c), these data indicate that some blood monocytes are dying of pyroptosis in COVID-19 infected patients.

### Circulating monocytes are infected with SARS-CoV-2 and the infected cells are undergoing pyroptosis

Pyroptosis in patient monocytes suggested that blood monocytes might take up SARS-CoV-2 to activate pyroptosis. Monocytes are not thought to express the receptor ACE-2, which serves as the dominant entry receptor for SARS-CoV-2, but this finding has been disputed (*38, 39*). Although we did not detect monocyte ACE-2 expression in HD monocytes, even after LPS activation, COVID-19 patient monocytes appeared to dimly express ACE-2 by flow cytometry and qRT-PCR (Extended Data Fig. 5a,b). Both HD and COVID-19 patient monocytes also expressed similar levels of CD147 (basigin or EMMPRIN), an immunoglobulin superfamily receptor implicated in bacterial, parasite and viral entry, which has been reported to bind to SARS-CoV-2 spike protein and facilitate viral uptake and infection (*40*). CD147 expression in HD monocytes did not change after LPS stimulation (Fig. S5c,d). Monocytes also express 2 Fc gamma receptors – CD32 (FcγRII, expressed on most blood monocytes (*41*)) and CD16 (FcγRIII, expressed on a small minority of blood monocytes that are activated and increased in number in COVID-19 patients (Fig. S1c)(*12*). These could recognize antibody-opsonized viral particles and mediate their entry via antibody-dependent phagocytosis (ADP)(*42*). Anti-SARS-CoV-2 spike protein antibodies are detected early in SARS-CoV-2 infection, about the same time as patients start developing inflammatory symptoms (*11, 43–45*). In fact, anti-Spike RBD antibodies were detected by ELISA in the plasma of most of the 18 COVID-19 patients we assayed at the time of presentation to the hospital ED (Fig. S6a). Two recent reports suggest that monocytes might be infected with SARS-CoV-2 (*15, 46*). To examine whether COVID-19 patient blood monocytes can be infected and support SARS-CoV-2 replication and to determine whether infection triggers inflammasome activation, we co-stained isolated HD and COVID-19 patient monocytes for SARS-CoV-2 nucleocapsid (N) (Fig 3e-h) or dsRNA (J2 antibody) (Fig 3i-l) and ASC. While cells that passively take up virions or contain replicating virus would be expected to stain for N, J2 staining generally indicates active viral replication (*47*). Although HD monocytes did not stain for N, dsRNA or ASC, ~10% of the blood monocytes from SARS-CoV-2 infected patients on average stained for N or dsRNA (Fig 3f,j), suggesting that a minority of circulating monocytes in COVID-19 patients contain actively replicating SARS-CoV-2. Moreover, virtually all of the infected cells showed ASC specks (Fig 3g,k) and virtually all of the cells with ASC specks were infected (Fig 3h,l). Thus SARS-CoV-2 infection of monocytes is associated with inflammasome activation and pyroptosis.

**Figure 3.**
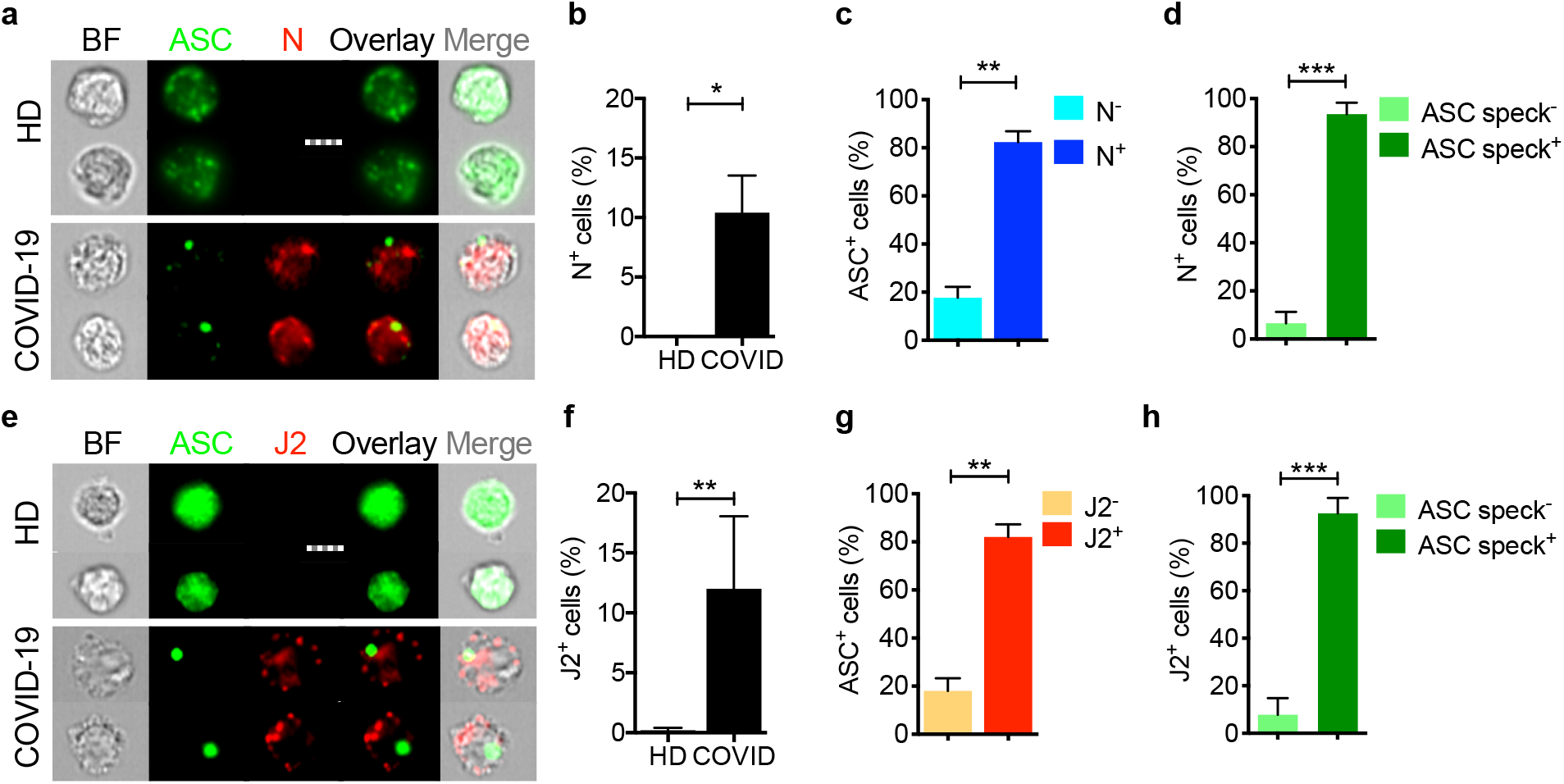
Circulating monocytes are infected with SARS-CoV-2. Circulating monocytes from HD and COVID-19 patients were purified and stained for SARS-CoV-2 nucleocapsid (N) (n=5) (**a-d**) or dsRNA (J2 antibody) (n=4) (**e-h**) and ASC. Shown are representative imaging flow cytometry images (**a**,**e**), quantification of percentage of cells that were infected by N (**b**) or J2 (**f**) staining, percentage of uninfected or infected cells (by J2 or N staining) that showed ASC specks (**c, g**) or percentage of cells with or without ASC specks that showed N (**d**) or J2 (**h**) staining. Scale bar, 7 μm. BF, brightfield. Mean ± S.E.M. is shown. *p<0.05, **p<0.01, ***p<0.001 by nonparametric unpaired *t* test (Mann-Whitney or Kolmogorov-Smirnov).

### Healthy donor monocytes are inefficiently infected with SARS-CoV-2, but infection is enhanced by anti-Spike antibody or patient plasma or by stimulation with LPS

To examine the susceptibility of monocytes to SARS-CoV-2 and begin to understand how they might become infected, purified HD monocytes were infected with an engineered infectious clone (icSARS-CoV-2-mNG) derived from the 2019-nCoV/USA_WA1/2020 strain and encoding a Neon Green (NG) fluorescent protein gene as a reporter of viral replication (*48*). Monocytes, primed or not with LPS, were infected (MOI 1) with the reporter virus that was preincubated with IgG1 isotype control antibody (mAb114), anti-Spike mAbs (non-neutralizing C1A-H12, neutralizing C1A-B12)(*49*) or pooled HD or COVID-19 patient plasma (heat inactivated or not). Antibodies and plasma were also present during the 48 h culture. After 48 h, monocytes were co-stained for dsRNA (J2 antibody) or SARS-CoV-2 N and ASC and analyzed by imaging flow cytometry (Fig. 4, Fig. S6b-i). Staining for N indicates virus internalization, whereas J2 staining and NG fluorescence indicate virus replication. Without LPS or anti-Spike antibody or COVID-19 pooled plasma, very few, if any, HD monocytes took up or replicated the virus, but infection (both uptake and replication) increased significantly in the presence of anti-Spike mAb or COVID-19 plasma. The neutralizing activity of the antibody did not appear to strongly affect infection. HD plasma did not augment HD monocyte infection and heat inactivation of COVID-19 plasma did not significantly alter its ability to potentiate infection. These findings suggested that complement did not facilitate monocyte infection and that specific antiviral antibodies that opsonize viral particles might be involved (Fig. S6b-i). Immunoglobulin (Ig)-depletion of COVID-19 plasma nearly abrogated viral infection assessed by J2 staining and NG fluorescence (Fig. 4k,l). Thus, viral entry in monocytes occurs predominantly by ADP of opsonized viral particles, but further work is needed to probe which Fc receptors are involved.

**Figure 4.**
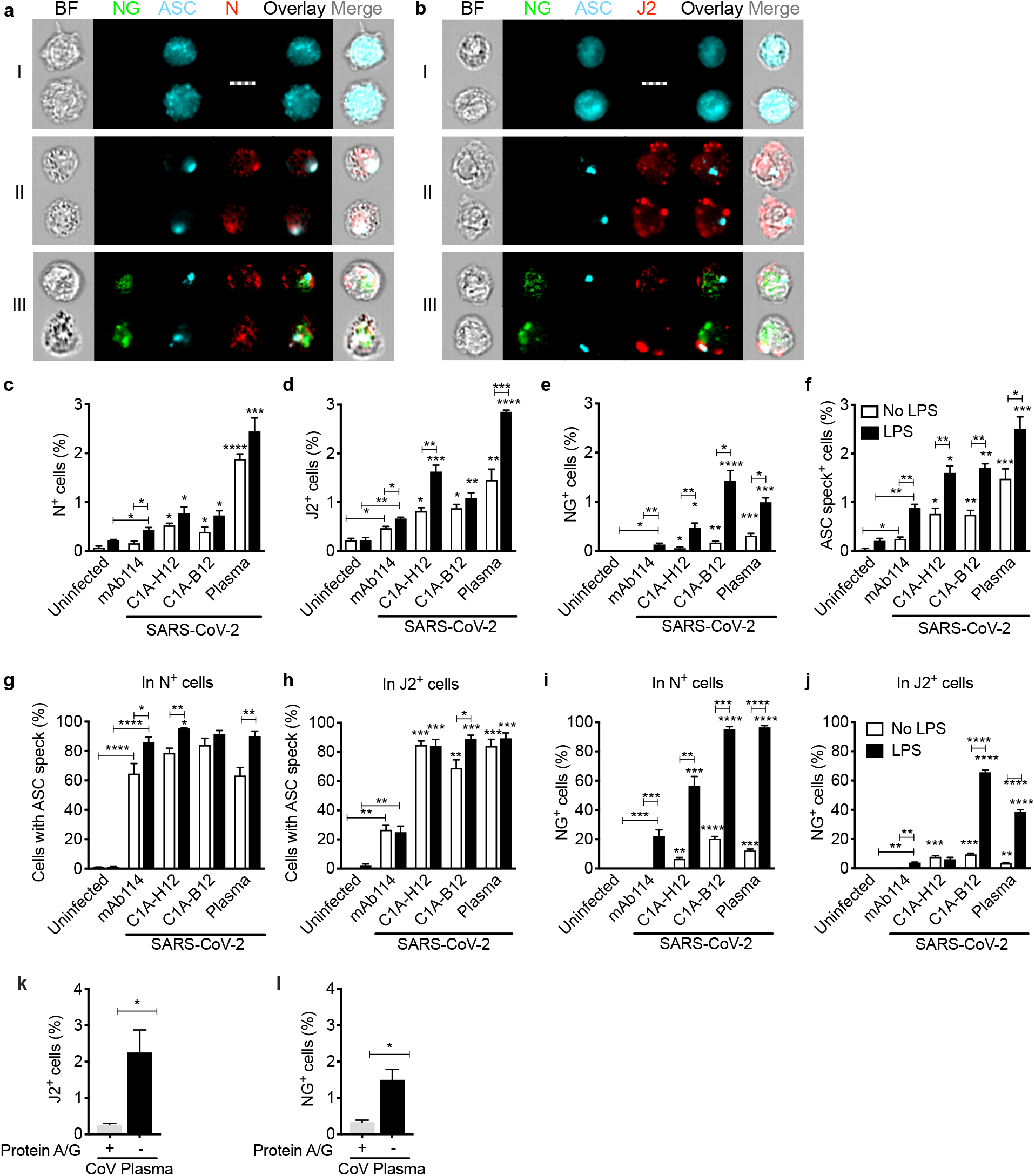
In vitro infection of healthy donor purified monocytes. Healthy donor (HD, n=3) monocytes were primed or not with LPS, infected with icSARS-CoV-2-mNG (MOI, 1), then stained 48 h later for nucleocapsid (N) or dsRNA (J2) and ASC. Before infection, virus was preincubated with indicated monoclonal antibodies (IgG1 isotype control mAb114), non-neutralizing anti-spike (C1A-H12) or neutralizing anti-RBD (C1A-B12) or with pooled COVID-19 patient plasma (Plasma). Antibodies or plasma were present throughout the culture. **a**,**b**, Representative imaging flow cytometry images of (I) uninfected monocytes, (II) N or J2 staining cells without detectable Neon green (NG), or (III) with NG and N or J2 staining. **c-f**, Quantification of HD monocytes staining for N (**c**), J2 (**d**), NG (**e**) or ASC specks (**f**). **g**,**h**, Percentage of N+ (**g**) and J2+ (**h**) cells that had ASC specks. **i**,**j**, Percentage of N+ (**i**) and J2+ (**j**) cells that had detectable NG. **k**,**l**, Percentage of J2+ (**k**) and NG fluorescent (**l**) LPS-activated monocytes after infection with icSARS-CoV-2-mNG virus that was preincubated with CoV pooled plasma that was pretreated with Protein A/G-coated agarose beads or with uncoated agarose beads to deplete or not immunoglobulins. BF, Brightfield. Scale bar, 7 μm. Mean ± S.E.M. is shown. *p<0.05, **p<0.01,***p<0.001, ****p<0.0001 relative to Iso or as indicated by two-way ANOVA with Sidak’s multiple comparisons test and unpaired non-parametric *t* test (k,l).

The highest in vitro infection rate was ~3% in HD monocytes pretreated with LPS and incubated with patient plasma. N and J2 staining were comparable across the different conditions with a low background of ~0.1% in uninfected samples; fewer cells were NG fluorescent (about half as many) and there was no background NG fluorescence in uninfected samples. More of the J2 or N staining cells in the samples with the highest infection rates (treated with LPS and patient plasma or anti-Spike antibodies) were also NG fluorescent, (Fig. 4i,j, Fig. S6g,h). We interpret these results to indicate that the virus replicates in most monocytes that take it up, but NG expression is detected only in cells with more vigorous replication. In particular LPS activation enhances SARS-CoV-2 infection and viral replication in monocytes. LPS pretreatment also slightly increased the frequency of J2 and N staining when anti-Spike or patient plasma were not added, suggesting that another non-Fc receptor on activated monocytes, such as ACE-2 or CD147, might weakly mediate viral uptake. Further work will be needed to identify how monocyte activation enhances monocyte infection in COVID-19 infection.

As in monocytes from COVID-19 patients, in vitro SARS-CoV-2 infection of HD monocytes led to ASC speck formation (Fig. 4a,b, f-h, Fig. S6). The numbers of infected HD monocytes with ASC specks under the various conditions (with and without antiviral antibodies or plasma, with and without LPS) roughly paralleled and were comparable to the numbers of anti-N or J2 staining cells. Furthermore, virtually all of the anti-N or J2 positive cells had ASC specks, suggesting that once a monocyte is infected with SARS-CoV-2, whether in COVID-19 patients in vivo or in ex vivo infection of HD monocytes, SARS-CoV-2 infection stimulates the formation of an inflammasome and pyroptosis.

## Discussion

Myeloid cells are sentinels that sound the innate immune alarm by sensing invasive infection and danger to activate inflammasomes. They are usually the most important source of inflammatory cytokines during inflammation. Processing and release of IL-1 family cytokines, probably the most potent inflammatory mediators in the body, depend on canonical inflammasome activation of caspase-1 and the pore-forming protein GSDMD (*8, 50*). Here we show that antibody-opsonized SARS-CoV-2 can infect and replicate in blood monocytes and that viral replication is enhanced by monocyte activation. As many as 10% of circulating monocytes in COVID-19 patients in our study were infected and a comparable number had activated inflammasomes and took up a small membrane-impermeable dye, indicating that they were undergoing pyroptosis. This is a very large number, considering that dying cells are usually difficult to detect in vivo since they are rapidly eliminated from the body. It may be surprising that monocyte infection and cell death has not been widely recognized. However, we think this is for three reasons – (1) many studies of COVID-19 blood cells use thawed, frozen cells, and dying or activated cells do not survive freeze-thawing, (2) published studies have not specifically looked at whether circulating mononuclear cells are dying and (3) few researchers have looked at whether monocytes might be infected because they were not thought to express ACE2. In support of our findings, a few studies have shown evidence of increased IL-1 family cytokines in COVID-19 patient plasma, in vitro SARS-CoV-2 entry in myeloid cells or NLRP3 inflammasome-caspase-1 activation in COVID-19 patient blood cells (*13–15, 46*). However, none of these studies has shown that SARS-CoV-2 infection of monocytes is antibody-mediated and leads to viral replication or that SARS-CoV-2 infection is so frequent in circulating monocytes. Moreover, the mechanism responsible for SARS-CoV-2 entry into monocytes and evidence of viral replication in monocytes has not been studied. Moreover, no previous study identified SARS-CoV-2 infection of monocytes as the cause of inflammasome activation or showed evidence of ongoing pyroptosis.

We found a one-to-one correspondence between monocyte infection and inflammasome-caspase-1 activation. SARS-CoV-2-infected monocytes had detectable NLRP3-ASC-caspase-1 and AIM-2-ASC-caspase-1 inflammasomes that recognize cell membrane damage and cytosolic DNA, respectively. How do these inflammasomes get activated by SARS-CoV-2 infection? Virulent coronaviruses express 3 ion channel proteins, Orf3a, Orf8 and E, that are attractive candidates, since they disrupt ion concentrations and when ectopically expressed have been shown to activate NLRP3 (*3, 26, 28, 30*). It is uncertain whether endogenous expression of these viroporins during native infection also activates NLRP3. We did not expect an RNA virus like SARS-CoV-2 to be sensed by the DNA sensor AIM2; further work is needed to determine whether the recognized DNA comes from the host or the virus. We also did not expect to find more than one inflammasome sensor stimulated in the same cell, but rare reports have previously noted co-localization of 2 distinct inflammasome sensors to the same speck (*51*). It will be worth studying whether other viral infections activate multiple inflammasomes, but as of now there are few studies of inflammasome activation and pyroptosis during any viral infection (*8*). The significant eQTL link of severe COVID disease to the *NLRC4* gene, which encodes an inflammasome that binds to the NAIP sensor of bacterial Type III secretion (*52*) hint that this inflammasome may play a role in severe COVID-19 disease, but if and how it does will need to be explored.

At the time of diagnosis plasma biomarkers of pyroptosis in our COVID-19 patient samples, including IL-1RA, IL-18, LDH and GSDMD, correlated with development of severe disease. This finding suggests that they might be incorporated into a diagnostic panel to help predict who might be susceptible to overexuberant inflammatory complications and benefit from immune modulating therapy. Repurposing FDA-approved drugs that inhibit inflammatory cytokines or GSDMD, the final mediator of both cytokine release and inflammatory death, are worth continuing to assess in controlled clinical trials. Although anti-IL-1β (canakinumab) did not meet its endpoint for efficacy in hospitalized hypoxic COVID-19 patients (Novartis press release, 11/06/2020), an unpublished manuscript of a randomized control trial of anakinra (IL-1RA) in patients with SARS-CoV-2 pneumonia showed a highly significant reduction in the development of severe respiratory failure and overall clinical severity (*53*). The possible efficacy of antagonizing IL-1 implicates GSDMD activation in severe COVID-19 disease since IL-1 secretion depends heavily on GSDMD pore formation. Antagonists of IL-6 signaling, however, have had weak, at best, effects on COVID-19 infection (*54, 55*). The disappointing results of inhibiting IL-6 may be due to suboptimal timing (it is hard to stop a fulminant inflammatory cascade once it has started) or because IL-6 is only one of many inflammatory mediators that are released and increased during severe disease. Two FDA-approved inhibitors of GSDMD, the critical mediator of pyroptosis and IL-1 family cytokine release - disulfiram (Antabuse) (*56*) or dimethyl fumarate (Tecfidera) (*57*) - are also worth evaluating. It is worth noting that administering disulfiram or dimethyl fumarate in mouse models of sepsis, which has many overlapping features with severe COVID-19 disease, strongly improved not only survival, but also plasma levels of IL-6 and TNFα.

In human studies like this, because of our inability to manipulate gene expression or deplete certain cell populations, it is difficult to assess how important monocyte infection and inflammasome activation is in COVID-19 inflammation, cytokine release syndrome and severe disease. However, given the large percentage of infected cells, the large number of monocytes in the blood (~1-3 ×10^9^) and the fact that myeloid cells are the major source of IL-1 and other inflammatory cytokines, it is likely that it is an important contributor. However, it will be worthwhile to study other possible infected cells that express GSDMD as sources of inflammation, especially pneumocytes and tissue resident macrophages in the lung and to understand what types of monocyte/macrophage activation enhance SARS-CoV-2 infection. The significant link we found of *GSDME* eQTLs to severe vs non-hospitalized COVID-19 infection even in a small case-control study (269 cases, 688 controls) suggests that it will also be worth identifying cell populations, infected or not, that have cleaved and activated GSDME in COVID-19 patients. GSDME-expressing cells switch from noninflammatory apoptosis to inflammatory pyroptosis, when they are exposed to apoptotic stimuli or to granzyme B during cytotoxic lymphocyte attack (*8*). Although there is no evidence that SARS-CoV-2 triggers effector caspase activation and apoptosis, cells infected with other virulent coronaviruses (SARS-CoV, MERS-CoV and mouse hepatitis virus (MHV)) have been shown to induce apoptosis (*3*).

Our findings implicate opsonizing antibodies in monocyte SARS-CoV-2 infection and inflammasome activation and inflammation and raise the possibility that antibody-dependent enhancement may be involved in deleterious immune reactions associated with severe disease (*58, 59*). If our hypothesis that pyroptotic inflammation underlies the development of severe disease is correct, it may not be a coincidence that clinical deterioration coincides temporally with the detection of SARS-CoV-2 antibody responses (*11, 43, 45*). Patients with severe COVID-19 disease have recently been shown to have a strong increase in antiviral IgGs that are afucosylated in their Fc region and bind more strongly to CD16, the main receptor that may be responsible for uptake of anti-Spike antibody-opsonized SARS-CoV-2 (*58, 60, 61*). Characterizing the antibodies that are most effective at enhancing monocyte infection will be important not only for understanding SARS-CoV-2 pathogenesis, but also for choosing the best preparations of convalescent patient plasma and monoclonal antibodies for therapy and/or prevention of severe disease and for comparing whether different vaccines generate antibodies that enhance monocyte infection and inflammation.

## Data Availability

Data and materials availability: The data and materials that support the findings of this study are available from the corresponding authors upon request.

## Acknowledgements

We thank members of the MGH COVID-19 collection and processing team (Kendall Lavin-Parsons, Brendan Lilley, Carl Lodenstein, Brenna McKaig, Nicole Charland, Hargun Khanna (Department of Emergency Medicine, MGH), Anna Gonye, Irena Gushterova, Tom Lasalle, Nihaarika Sharma (MGH Cancer Center), Brian C. Russo, Maricarmen Rojas-Lopez (Division of Infectious Diseases, Department of Medicine, MGH) and Moshe Sade-Feldman, Kasidet Manakongtreecheep, Jessica Tantivit, Molly Fisher Thomas (MGH Center for Immunology and Inflammatory Diseases) for plasma samples. We also thank the Analytical Instrumentation Core Lab of Boston University for running and analyzing the Luminex Multiplex assay.

## Funding

This research was supported by:

National Institutes of Health grant R01AI124491 (HW)

National Institutes of Health grant U19AI131135 (LG)

Annenberg Foundation and FAST Grants and a gift from Jeanne Sullivan (AEG)

American Lung Association (MBG, MRF)

British Heart Foundation Programme Grant RG/16/4/32218 (SB)

Conselho Nacional de Desenvolvimento Científico e Tecnológico (CNPq) fellowship (CJ)

## Author Contributions

All authors contributed to manuscript preparation.

Conceptualization: JL, CJ, AC, HW

Experimentation: CJ, AC, SR, JI, FH, SH, LBdeL, ML, SV, LH, EJ, VL, BP, GM, SB

Patient recruitment: CB, JM, NR, UDA, DK, MF, MG Data analysis: CJ, AC, JI, SB, CS

Reagents: SC, JA

Supervision: JL, CJ, AC, LG, AG

Manuscript writing: AC, CJ, JI, JL

## Competing Interests

The authors declare no competing interests.

### Data and materials availability

The data and materials that support the findings of this study are available from the corresponding authors upon request.

## Supplementary Information is available for this paper

**Supplemental Table S1: Demographic and clinical information of the fresh PBMCs and plasma cohort**. Age, race and ethnicity, body mass index, co-morbidities, symptoms, MGH Acuity score, hospitalization details and clinical information of the patients in the fresh PBMCs and plasma cohort.

**Supplemental Table S2: Demographic and clinical information of the frozen plasma cohort**. Age, body mass index, co-morbidities, symptoms, MGH Acuity score,hospitalization details and clinical information of the patients in the frozen plasma cohort.

**Supplemental Table S3: eQTL data**

**Supplemental Table S4: Reagents and materials (with respective sources) used for this manuscript**. Antibodies, chemicals and commercial kits (with sources and catalog numbers) described in Methods.

## Methods

### Human subjects

#### Fresh PBMCs and plasma cohort

The study was approved by the Investigation Review Boards of Boston Children’s Hospital and Massachusetts General Hospital (MGH), and all enrolled patients signed an informed consent. 35 patients 18 years or older with clinical symptoms suggestive of COVID-19 infection were enrolled at the time of presentation to the MGH emergency department (ED) from 7/9/20 to 01/12/21. A 10-ml EDTA blood sample was transported to Boston Children’s Hospital and processed within 2 h of collection. Only samples from patients with qRT-PCR verified SARS-CoV-2 infection were included in the study (31). Demographic and clinical data are summarized in Table S1. Healthy donor (HD) samples were processed and analyzed in parallel with patient samples.

#### *Frozen plasma cohort* 60 patients

18 yr or older with clinical symptoms suggestive of COVID-19 infection were enrolled in the MGH ED from 3/15/20 to 4/15/20 with an IRB-approved waiver of informed consent. Enrolled patients had at least one of the following: (i) tachypnea ≥22 breaths per minute, (ii) oxygen saturation ≤92% on room air, (iii) requirement for supplemental oxygen, or (iv) positive-pressure ventilation. A 10-ml EDTA tube was obtained with the initial clinical blood draw in the ED (n=60). Blood was also obtained on days 3 (n=42) and 7 (n=35) if the patient was hospitalized on those dates. Clinical course was followed for 28 d post-enrollment or until hospital discharge if after 28 d. SARS-CoV-2-confirmed patients (by qRT-PCR) were assigned a maximum acuity score (A1-A5) (A1 – died, A2 – required mechanical ventilation, A3 – hospitalized requiring supplemental oxygen, A4 – hospitalized but not requiring supplemental oxygen, A5 – discharged and not requiring hospitalization)(*20, 21*). Patients were grouped based on their worst acuity score over 28 d and divided into three groups for comparison (A1 and A2, severe disease; A3, moderate disease; A4 and A5, mild disease). Only 1 patient was in A4; therefore, most mild patients represent those that were discharged directly from the ED and thus have only a day 0 sample. CRP and D-dimer values were grouped in the following categories: CRP (mg/L) 1 = 0-19.9, 2 = 20-59.0, 3 = 60-99.9, 4 = 100-179, 5 = 180+; D-dimer (ng/mL) 1 = 0-499, 2 = 500-999, 3= 1000-1999, 4 = 2000-3999, 5 = 4000+. Demographic and clinical data are summarized for each outcome group (Table S2).

### Plasma, PBMC and monocyte isolation

Samples were processed using recommended safety precautions in a BSL-2+ facility. Blood tubes were centrifuged at 2000 rpm for 10 min to separate plasma from blood cells. Plasma was collected to a new tube and incubated or not with 1% Triton X-100 for 1 h on ice before aliquoting and freezing at −80°C. Blood cells were resuspended in PBS and layered over Ficoll for density centrifugation. PBMC were collected from the interface and subjected to red blood cell lysis (if necessary) with Red Blood Cell Lysing Buffer Hybri-Max for 5 min on ice, followed by quenching with RPMI medium supplemented with 10% FBS and 1% Penicillin/Streptomycin. PBMC were washed once more with RPMI and one fraction was stained for flow cytometry, while the remaining cells were used for monocyte purification by magnetic separation using CD14+ magnetic beads. Sources of reagents are described in Table S3.

### Multiplex Luminex, ELISA and LDH activity assay

IL-1β, IL-1RA, IL-2, IL-4, IL-5, IL-6, IL-7, IL-10, IL-12, IL-13, IL-17, IL-18, IL-21, IL-23, CCL3, CCL7, CCL9, CXCL10, G-CSF, TNF-α, IFN-β and IFN-γ were measured in plasma samples using a custom Luminex assay (R&D Systems), following the manufacturer’s instructions. Plates were analyzed using a Luminex MAGPIX Analyzer at the Analytical Instrumentation Core Lab of Boston University. GSDMD was measured in the same samples using the Human GSDMD ELISA kit (MyBiosource) following the manufacturer’s instructions, and LDH activity was measured using the CytoTox 96® Non-Radioactive Cytotoxicity Assay (Promega). Results from the latter assays were analyzed using a Biotek Synergy 2 analyzer; GSDMD absorbance was measured at 450 nm and LDH absorbance was measured at 490 nm.

### Monocyte treatment and culture

Purified monocytes and THP-1 cells (ATCC) as controls, were cultured in RPMI + 10% FBS + 1% Pen/Strep for 30 min at 37°C in the presence or absence of 20 µM nigericin. Following incubation, monocytes were washed and used for FLICA assay, or fixed for 10 min with 4% PFA, washed twice with PBS + 3% FBS and kept at 4° C in PBS + 3% FBS until imaging flow cytometry or confocal microscopy.

As a positive control for pyrin activation, HD monocytes were cultured overnight with 2 µg/ml C3 Transferase from Clostridium botulinum (Rho I inhibitor), then fixed for 10 min with 4% PFA before staining for imaging flow cytometry (data not shown).

### Intracellular staining for imaging flow cytometry and confocal microscopy

Fixed monocytes were permeabilized with 0.1% Triton X-100 for 10 min and washed twice with PBS + 3% FBS. Monocytes were then blocked for 30 min with PBS + 5% FBS, washed twice and then stained with unconjugated primary antibodies for ASC (1:200, mouse or rabbit), NLRP3 (1:200, goat), AIM2 (1:200, mouse), GSDMD (1:200, mouse), pyrin (1:200, rabbit), dsRNA (J2, mouse) (1:500) or SARS-CoV-2 nucleocapsid protein (1:500, rabbit) for 2 h, followed by 3 washes with PBS + 3% FBS. Cells were then stained with secondary antibodies (donkey anti-mouse, rabbit or goat conjugated with AlexaFluor 488, 546 or 647, at 1:1000) for 1 h in PBS + 3% FBS, followed by 3 washes.

For microscopy, cells were then stained with DAPI (1:1000) for 10 min, washed 3 times and cytospun onto glass slides (VWR); sealed using polyvinyl alcohol and 1.5 mm coverslips (VWR). Confocal images were acquired using a Zeiss LSM 800 with 405, 488, 561 and 633 nm lasers (emission filters, 465, 509, 561 and 668 nm, respectively) and a 63x 1.4NA oil immersion objective. Images were processed using Zen Blue 3.2.

For imaging flow cytometry, cells were resuspended in PBS + 3% FBS for analysis. Data were acquired using an ImageStream X MKII (Amnis) with 63x magnification and analyzed using Ideas software (Amnis). Monocytes were gated on area/aspect ratio. ASC, NLRP3, AIM2 and Pyrin specks were gated and quantified based on fluorophore intensity/max pixels.

### Flow cytometry

PBMC were washed and stained for viability with Zombie Yellow in PBS (1:200) for 15 min on ice. Cells were washed with PBS, centrifuged, and then stained with Annexin V PE (1:200) in 1x Annexin Buffer for 15 min on ice. After washing with 1x Annexin V buffer, cells were blocked for 10 min with anti-CD32 (1:100) in PBS + 3% FBS, and then stained for 15 min on ice with a cocktail of antibodies to identify lymphocyte and myeloid cell subsets (all 1:200 except CD19 BV650, CD123 PerCP-Cy5.5 and CD56 APC-Cy7, 1:100). Purified monocytes and an A549 cell line overexpressing ACE2 were blocked with anti-CD32, then stained with primary antibodies for ACE2 (1:100) for 15 min on ice. The secondary anti-goat AF488 was coincubated with CD14 PE-Cy7 (1:200) and CD147 APC (1:100). After the last wash, cells were resuspended in 2% PFA and kept at 4°C until flow cytometry analysis. In vitro-infected monocytes were fixed and permeabilized with 0.1% Triton X-100, then blocked with PBS + 5% FBS. Cells were stained with primary antibodies for dsRNA (J2, mouse) (1:500), then stained with secondary antibody (donkey anti-mouse conjugated with AlexaFluor 647, at 1:500) and anti-CD14 PE-Cy7. Cells were acquired using a FACS Canto II or LSR II and data were analyzed using FlowJo Version 10.

### FLICA assay

Monocytes, cultured or not for 30 min with nigericin, were washed and resuspended in RPMI 10% FBS with FLICA substrate (BioRad FAM-FLICA Caspase-1 kit), and cultured for 1 h at 37° C. Cells were then washed twice with 1X Apoptosis Buffer (from the kit) and fixed with 1x Fixative (from the kit). Cells were kept at 4°C until further staining and analysis.

### Immunoblot

Lysates of enriched monocytes from HD and COVID-19 patients, the former treated or not for 16 h at 37°C with 100 ng/ml LPS and 20 μM nigericin, were resolved on 12% SDS PAGE gels, transferred to nitrocellulose membranes and blotted to detect GSDMD using (Abcam ab210070) primary rabbit mAb and secondary anti-rabbit IgG. Blotting for β-actin and COX-IV were used as loading controls.

### eQTL analysis

To assess whether a causal association exists between *GSDMD* and other immune gene eQTLs and severe COVID-19, *in silico* analyses were performed using two sample Mendelian randomization (*62*) in R v4.0.2 (*63*) using the *TwoSampleMR* package (*64*). Mendelian randomization is a form of instrumental variable analysis that exploits the random allocation of alleles at meiosis to draw causal inferences using observational data by attempting to emulate randomization procedures that would be adopted in a clinical trial.

Uncorrelated single-nucleotide polymorphisms (SNPs) (r^2^ < 0.001 in European ancestry individuals in the 1000 Genomes Project, Phase 3 release) were associated with whole-blood RNA expression of *GSDMD* and other immune genes at genome-wide significance (P<5×10^−8^) from the eQTLGen consortium (*22*). These SNPs were cross-referenced against a large phenotypic database of publicly available genetic associations to ensure that they were not associated with potential confounding factors (*65, 66*). Summary statistics from a genome-wide association study (*67*) of severe COVID-19 with respiratory failure were used for outcome data (*23*). Analysis was performed with data from release #4 (October, 2020) in which there were 4336 severe COVID-19 patients versus 623,902 control subjects, 6406 hospitalized COVID-19 patients versus 902,088 control subjects, and 269 severe COVID-19 patients vs 688 hospitalized COVID-19 patients. These analyses were based on different sample sets depending on whether the original investigators collected the relevant information either when planning the study or were able to obtain it retrospectively. In particular 14 studies contributed data to “COVID-19 hospitalized patients vs control population” and 12 studies contributed to “severe COVID-19 patients vs control population”. The control populations were a mix of subjects who were not COVID-19 infected (ie, negative test result(s)) or were assumed to be not COVID-19 infected (ie, there was no record of Covid-19 in their linked data).

CrossMap (*68*) was used to convert genomic positions from hg38 (as reported in the COVID-19 GWAS) to hg19 using the UCSC liftover chain file to ensure both the exposure and outcome datasets were reported on the same genome assembly. Variants were aligned so that the effect alleles were consistent across studies. The proportion of variance in expression of selected immune genes in whole blood explained by the selected SNPs the expected *F* value to examine potential weak instrument bias were then calculated (*69*).

Our primary analysis was based on the inverse variance weighted method of performing Mendelian randomization (this method combines the causal effect estimates from each individual genetic variant, computed as the ratio of the variant-expression association to the variant-Covid-19 association, into a single causal effect). A range of sensitivity analyses were performed relaxing some of the stricter assumptions underlying this method including the weighted median, modal and MR-Egger methods (*70*). If only a single genetic variant was selected for a gene, the Wald ratio method was used. The expected *F* value for *GSDMD* was 504.9 (with a lower limit of the one-sided 95% confidence interval (95% CI) of 462.1 indicating that considerable weak instrument bias would not be expected). There was no evidence of vertical pleiotropy (MR-Egger intercept p-value = 0.99) and findings were consistent across all sensitivity analyses.

Effect estimates are presented as odds ratios per standard deviation increase in *GSDMD* expression. A p-value < 0.05 was considered significant. All summary data used in this work are publicly available, together with a description of relevant participant consent and ethical approval secured in the original investigation.

### In vitro SARS-CoV-2 infection of HD monocytes

icSARS-CoV-2-mNG (a molecular clone of SARS-CoV-2 expressing Neon Green fluorescent protein) was a gift to AEG from Shi Pei Yong and the World Reference Center for Emerging Viruses and Arboviruses, Department of Microbiology and Immunology, University of Texas Medical Branch, Galveston, TX)(*48*). The NG fusion protein is only expressed during viral replication. HD monocytes (purified from apheresis leukoreduction collars collected at Brigham and Women’s Hospital) were incubated overnight with medium or 100 ng/ml LPS, and then infected with icSARS-CoV-2-mNG (MOI =1) in a BSL-3 facility. The viral inoculum was treated with 10 μg/ml of antibody (isotype control mAb114, anti-Spike C1A-H12, or anti-Spike C1A-B12), or 10% HD or COVID-19 patient pooled plasma (heat inactivated or not; Ig-depleted or not, as indicated) before infection with SARS-CoV-2 for 30 min at room temperature. 100 μl of treated virus was added to monocytes (2×10^6^ cells/well) in 48 well plates. Infected cells were incubated at 37°C, 5% CO_2_ with gentle shaking every 10 min for 1 h, after which the culture volume was increased to 500 μl with RPMI supplemented with 5% heat inactivated normal AB human serum and 10 μg/ml of the aforementioned antibodies or 10% pooled HD or COVID-19 patient plasma. Cultures were then incubated at 37°C, 5% CO_2_ for 48 h at which time cells were harvested and fixed for 20 min with 4% PFA and then stained. Immunoglobulin (Ig) from COVID-19 patient pooled plasma was depleted by protein A/G agarose resin. Control samples were incubated with agarose resin without coupled protein. C1A-B12 and C1A-H12, two SARS-CoV-2 Spike-targeting human monoclonal antibodies, were produced as previously described (*49*).

### qRT-PCR

RNA extracted from HD monocytes (stimulated or not with LPS (100 ng/ml for 16 h), or COVID-19 patient monocytes was reverse transcribed using a High Capacity cDNA Reverse Transcription Kit (Applied Biosystems) and the cDNA was analyzed by qRT-PCR using the Sso Fast™ EvaGreen® Supermix (BioRad) (30 sec at 95°C, 40 cycles (3 sec at 95°C; 3 sec at 54 °C) for both *ACE2* and *BSG* (CD147)) using a CFX96 Touch Real-Time PCR Detection System (BioRad). Primer sequences for *ACE2, BSG* and *ACTB* are given in Table S4.

### Anti-Spike RBD ELISA

Enzyme-linked Immunosorbent Assay (ELISA) kit anti-Spike RBD (BioLegend) was used to quantify antigen-specific IgG in plasma from HD and COVID-19 patients. ELISA was performed as per manufacturer’s instructions. Anti-Spike RBD absorbance was measured at 450 nm and quantified by linear regression based on the standard curve.

### Statistical Analysis

Statistical analysis was performed using GraphPad Prism V7.0. Normal distribution of the data was evaluated by the D’Agostino and Pearson normality test prior to applying statistical methods. Distributions were considered normal if *P* ≤ 0.05. Parametric or non-parametric (Mann-Whitney or Kolmogorov-Smirnov tests) two-tailed unpaired *t*-tests were used to compare two unpaired groups. Multiple group comparisons were analyzed by one-way ANOVA with Sidak’s or Tukey’s multiple comparisons tests, or non-parametric Kruskal-Wallis with Dunn’s post-test. Multiple groups were compared by two-way ANOVA with additional Sidak’s or Tukey’s multiple comparisons test. Mean plasma values from hospitalized COVID-19 patients on each day were compared between severity groups by multiple unpaired *t*-tests. Correlations of plasma levels were determined by simple linear regression and Pearson correlation coefficient. Differences were considered statistically significant when *P* ≤ 0.05.

## Notes

### Competing Interest Statement

The authors have declared no competing interest.

### Author Declarations

Fresh PBMCs and plasma cohort. The study was approved by the Investigation Review Boards of Boston Children's Hospital and Massachusetts General Hospital (MGH) and all enrolled patients signed an informed consent. Frozen plasma cohort. 60 patients 18 yr or older with clinical symptoms suggestive of COVID-19 infection were enrolled in the MGH ED from 3/15/20 to 4/15/20 with an IRB-approved waiver of informed consent.

