## Supplementary figure for "SARS-CoV-2 infects blood monocytes to activate NLRP3 and AIM2 inflammasomes, pyroptosis and cytokine release"

### Supplementary data

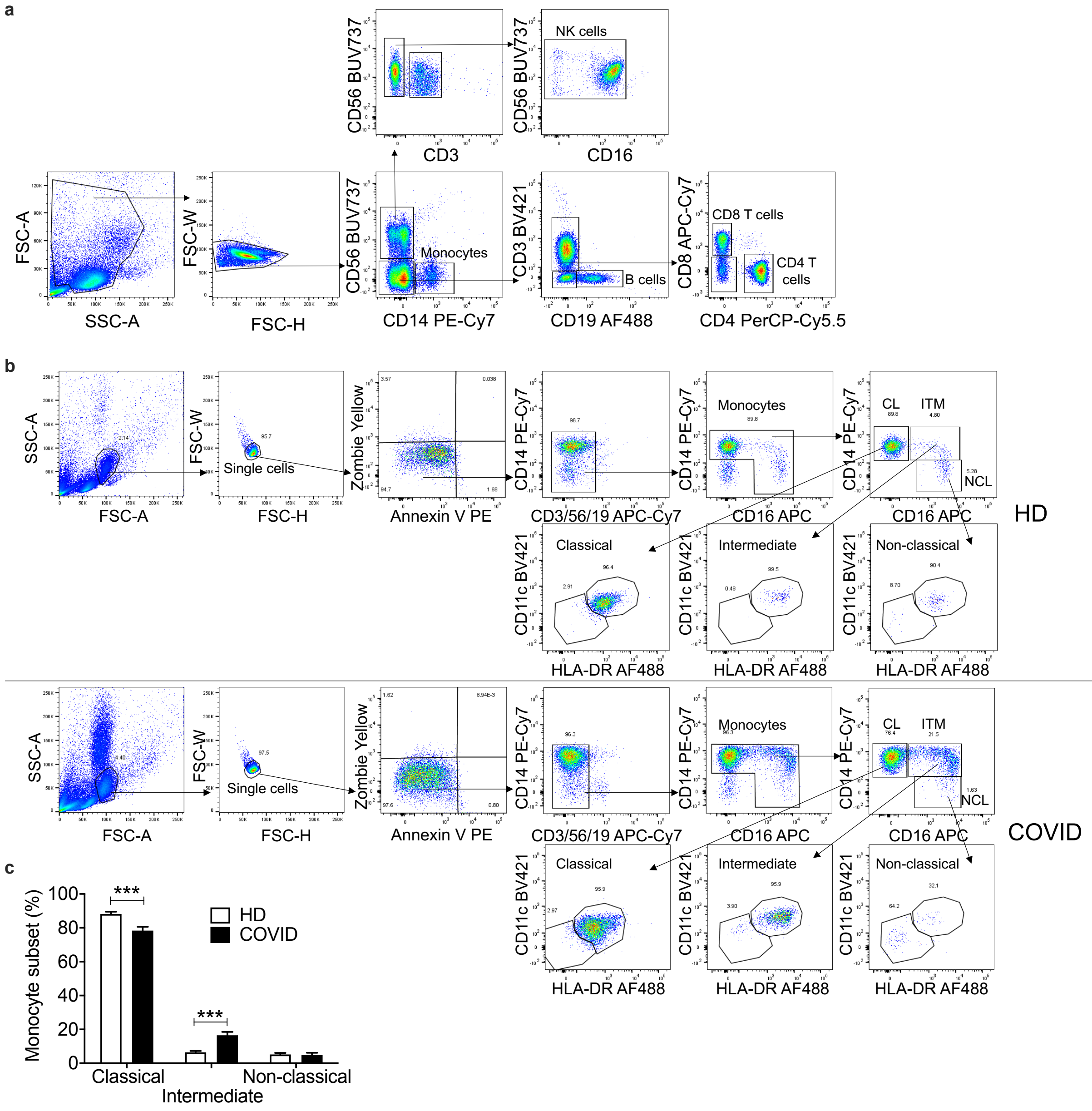


**Figure S1. Identification of lymphocyte and monocyte subsets in healthy donors and COVID-19 patients. a,** Gating strategy for identifying lymphocytes and monocytes in Figure 1. **b,** Gating strategy to identify monocyte subsets (classical (CL), intermediate (ITM) and non-classical (NCL)). **c,** Percentages of monocyte subsets in HD (n=10) and COVID-19 (n=12) patients. Bars represent mean ± S.E.M. ***$p < 0.001$ by two-way ANOVA followed by Sidak multiple comparisons test.

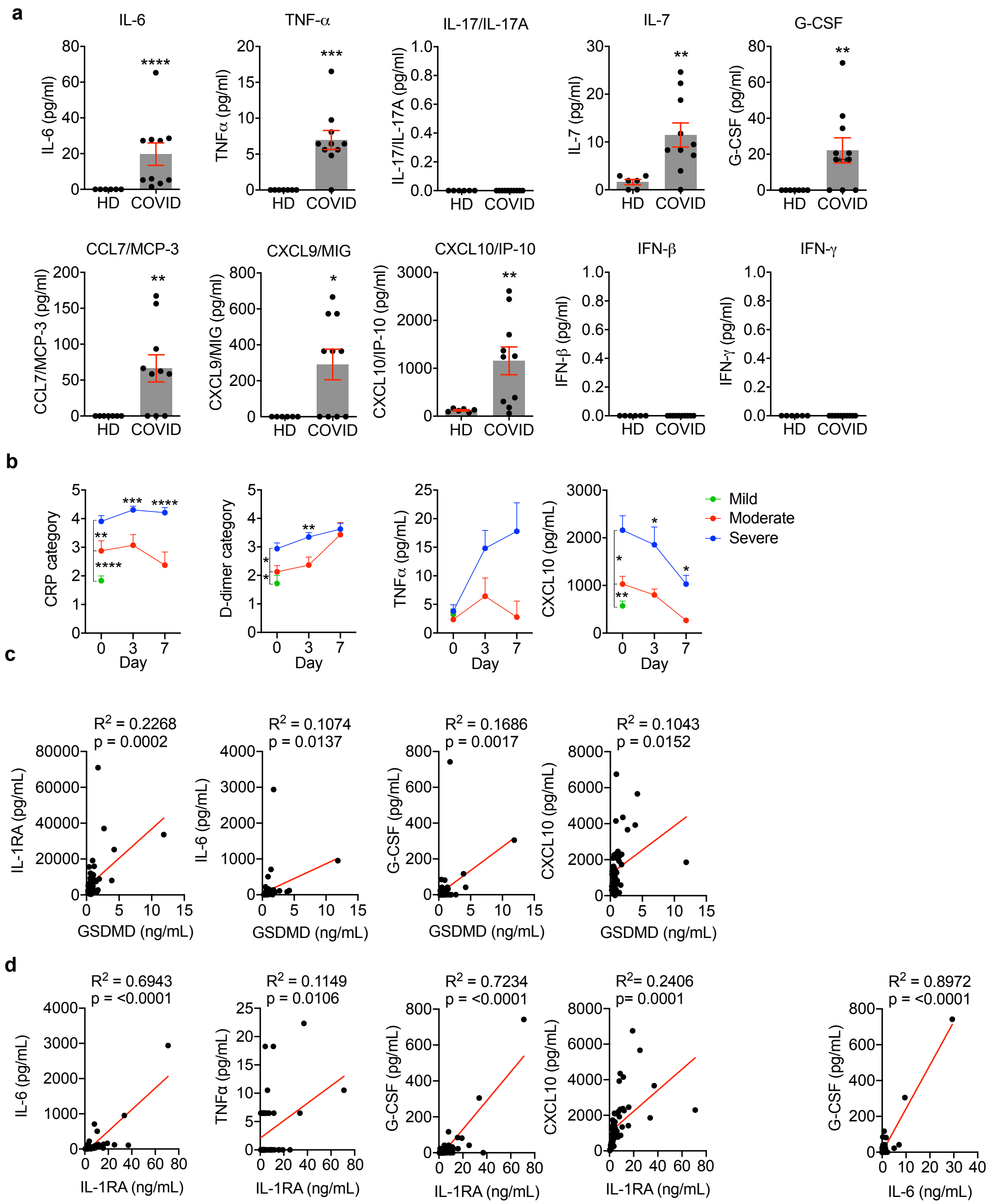


**Figure S2. COVID-19 patient plasma shows evidence of increased inflammation a,** Concentration of inflammatory cytokines (IL-6, TNF-α, IL-17/IL-17A), growth factors (IL-7, G-CSF), chemokines (CCL7, CCL9, CXCL10) and interferons (IFN-β, IFNγ) in the plasma of healthy donors (HD, n=6) or COVID-19 patients (n=10). **b,** Plasma C-reactive protein (CRP), D-dimer, TNF-α and CXCL10 in mild, moderate and severe COVID-19 patients (n=60) over time during hospitalization. CRP and D-dimer values were grouped as described in the methods. **c,** Correlation of GSDMD levels in the plasma of COVID-19 patients (at time of diagnosis, day 0) with IL-1RA, IL-6, G-CSF, and CXCL10. **d**, Correlation of day 0 plasma IL-1RA with IL-6, TNFα, G-CSF and CXCL10, and of IL-6 with G-CSF. Data shown are mean ± S.E.M. *p<0.05, **p<0.01, *** p<0.001 (by Mann-Whitney or Kolmogorov-Smirnov test (**a**), multiple t-tests (**b**) and simple linear regression and Pearson correlation (**c,d**)).

**a**

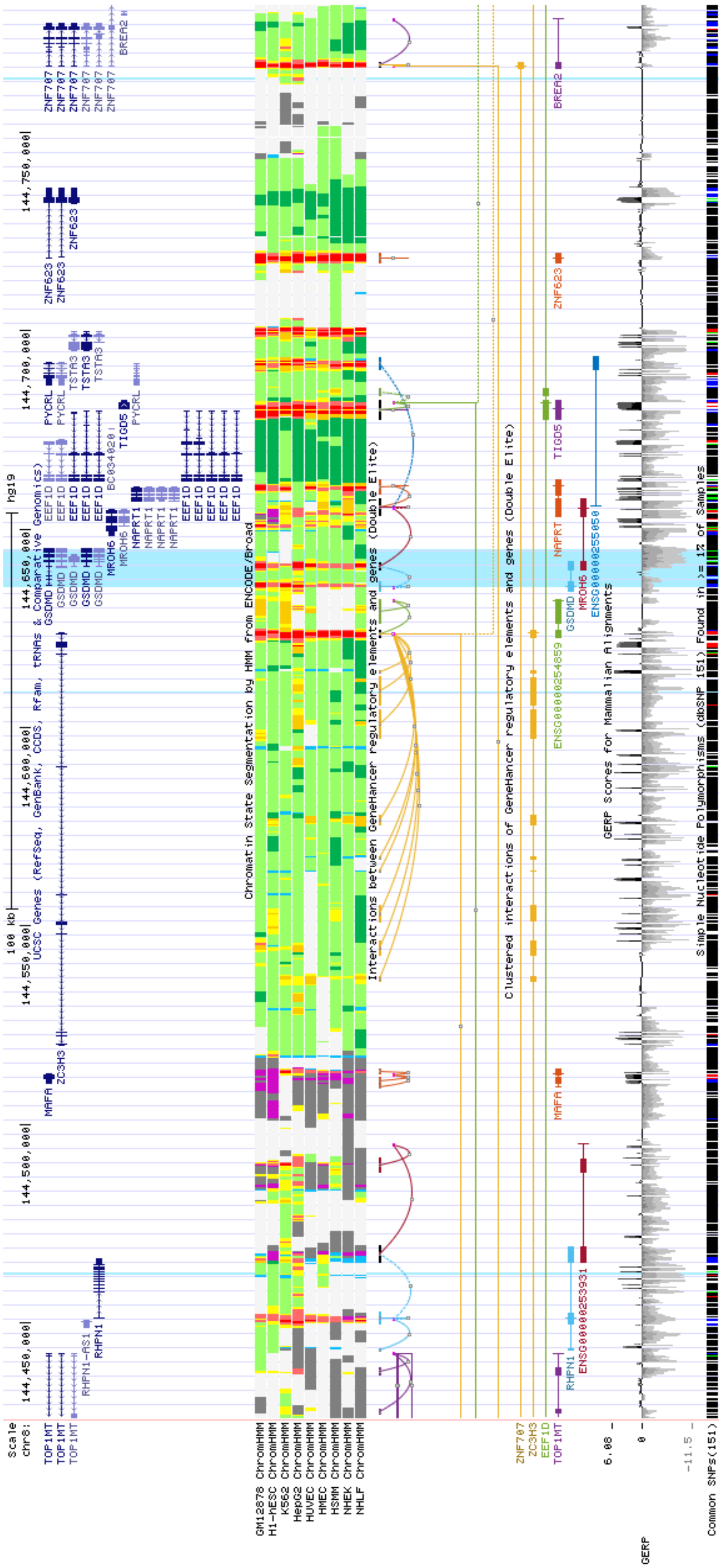

**b** **c**

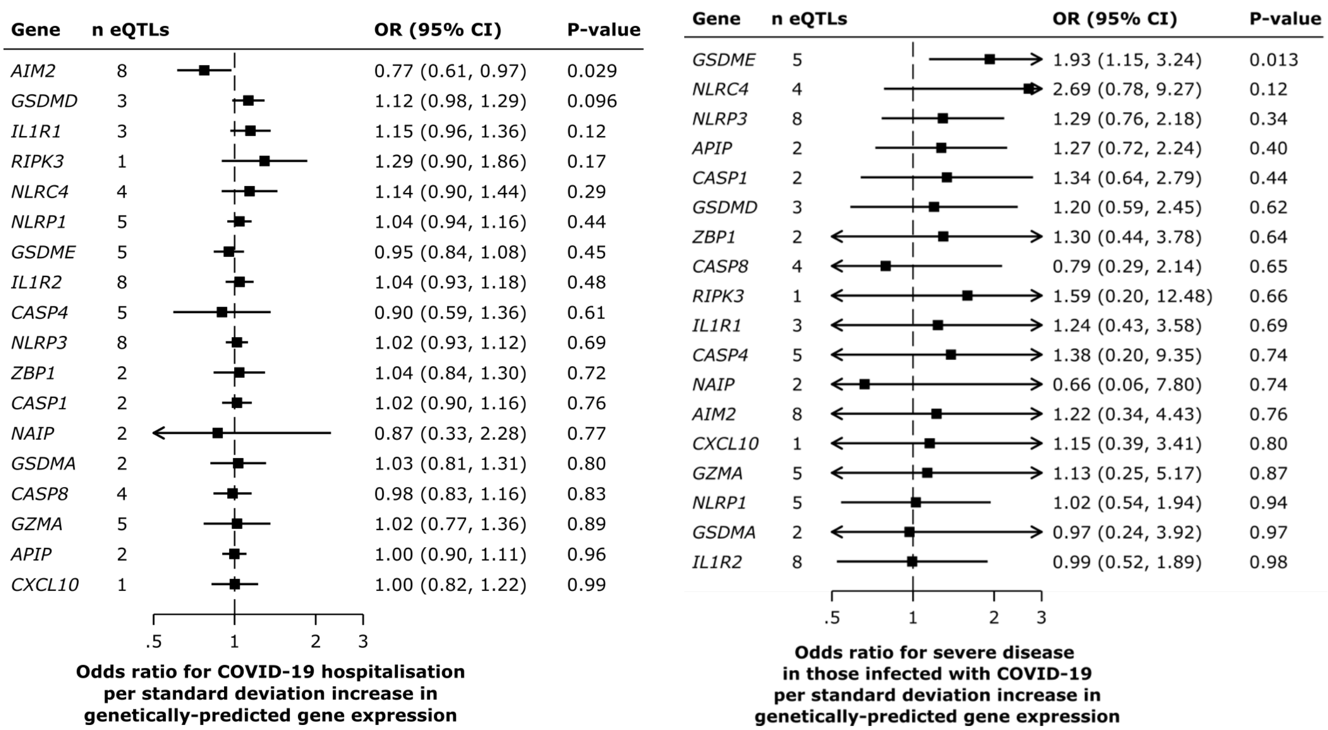


**Figure S3. Analysis of genetic link of immune gene eQTLs to severe COVID-19 disease a,** Screenshot of UCSC genome browser in the vicinity of *GSDMD* (highlighted in turquoise) on chromosome 8. The eQTLs are the thin turquoise vertical lines. None are within the *GSDMD* gene - one is within the gene *RHPN1*, one is within *ZC3H3*, and one is adjacent to *ZNF707*. All are eQTLs (validated by the eQTLGen consortium)[21] that are associated with increased *GSDMD* expression. **b,** Analysis of eQTL links to COVID-19 hospitalization (6406 hospitalized cases versus 902,088 population controls), **c,** Analysis of eQTL links in patients with severe COVID-19 disease (269 cases) vs control infected patients who did not require hospitalization for COVID-19 (688 controls).

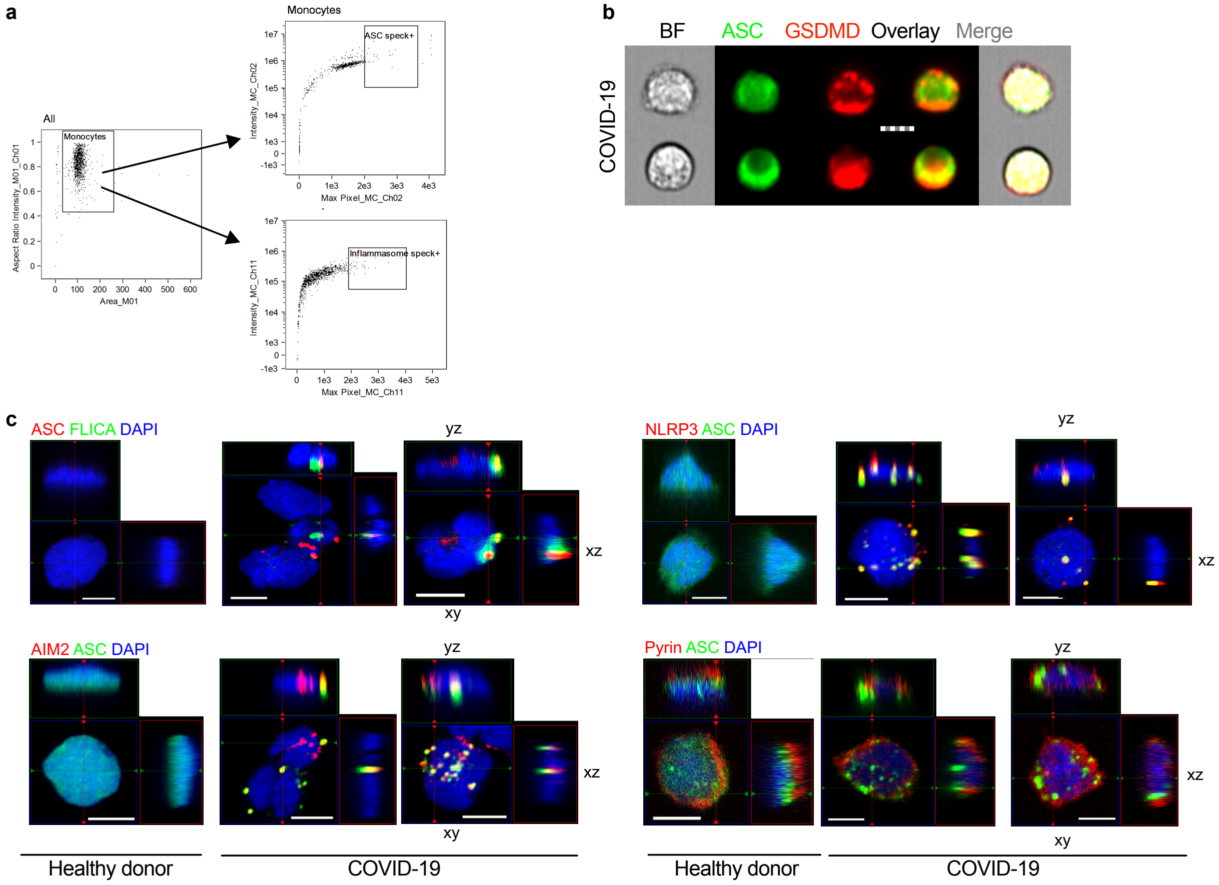


**Figure S4. a,** Gating strategy for imaging flow cytometry analysis of isolated monocytes. **b,** Representative imaging flow cytometry images of GSDMD and ASC co-staining in COVID-19 patient monocytes that lacked ASC specks. Scale bar, 7 µm. **c,** Representative confocal image z-stacks and plane projections of monocytes of HD and COVID-19 patient monocytes, stained for the same markers as in Figure 2. Scale bars, 5 µm.

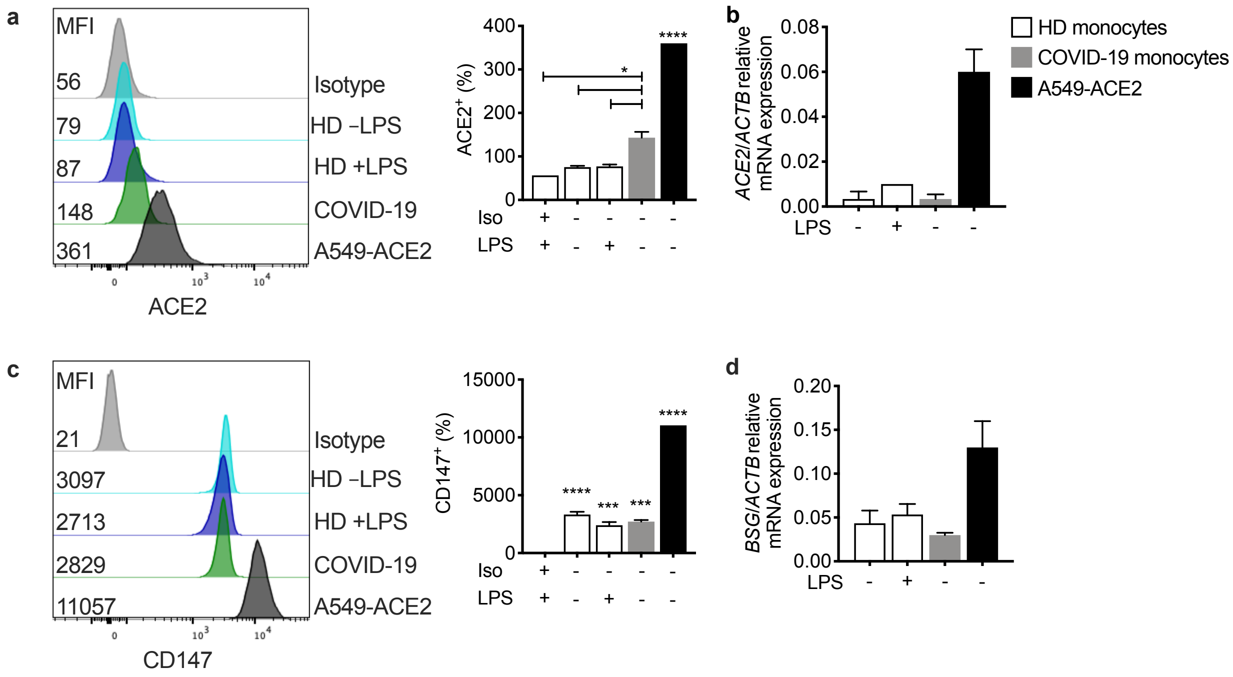


**Figure S5. ACE2 and CD147 expression on circulating monocytes.** Purified blood monocytes from HD (n=3) and COVID-19 patients (n=4) were analyzed by flow cytometry (**a,c**) or qRT-PCR (**b,d**) for expression of ACE2 (**a,b**) or CD147 (*BSG*) (**c,d**). HD monocytes were treated or not with LPS before analysis. A549-ACE2 cells were used as positive control. Mean ± S.E.M. is shown. *p<0.05, **p<0.01, ***p<0.001, ****p<0.0001 relative to isotype control-stained LPS activated HD monocytes (a,c) by one-way ANOVA with Tukey's multiple comparisons test.

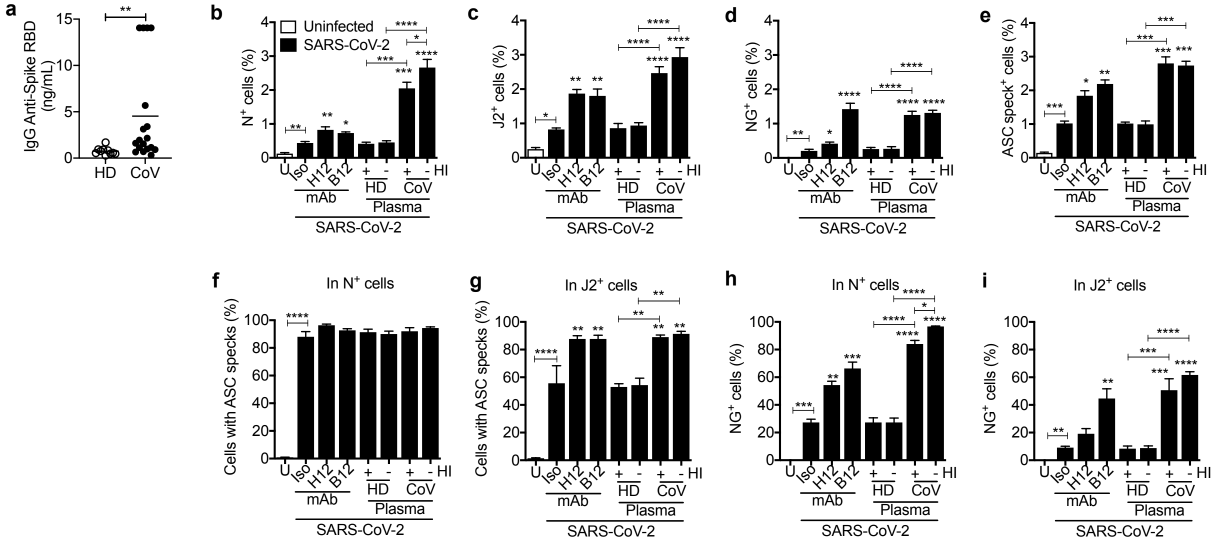


**Figure S6. Effect of anti-spike monoclonal antibodies or pooled COVID plasma on in vitro infection of LPS-activated healthy donor purified monocytes with icSARS-CoV-2-mNG a**, Spike RBD-specific IgG in the plasma of healthy donor (HD=10) and COVID-19 patient plasma (n=18) at the time of presentation in the ED was quantified by ELISA. **b-i**, HD monocytes were primed with LPS, infected with icSARS-CoV-2-mNG (MOI, 1), then stained 48 h later for nucleocapsid (N) or dsRNA (J2) and ASC and analyzed by imaging flow cytometry. Before infection, virus was preincubated with indicated monoclonal antibodies (IgG1 isotype control mAb114 (Iso)), non-neutralizing anti-spike (C1A-H12 (H12)) or neutralizing anti-RBD (C1A-B12 (B12)) or with pooled HD or COVID-19 patient plasma that had been heat-inactivated (HI) or not. U, uninfected. Quantification of HD monocyte staining for N (**b**), J2 (**c**), NG (**d**) or ASC specks (**e**). **f,g**, Percentage of N+ (**f**) and J2+ (**g**) cells that had ASC specks. **h,i**, Percentage of N+ (**h**) and J2+ (**i**) cells that had detectable NG. Mean ± S.E.M. is shown. *$p<0.05$, **$p<0.01$, ***$p<0.001$, ****$p<0.0001$ relative to Iso or as indicated, by two-way ANOVA with Sidak's multiple comparisons test.
